# Influenza vaccination coverage among healthcare workers during the first two seasons of data collection, Finland, 2017-2019

**DOI:** 10.1101/2020.09.08.20174771

**Authors:** Charlotte C Hammer, Outi Lyytikäinen, Dinah Arifulla, Saija Toura, Hanna Nohynek

## Abstract

**Background:** Influenza can cause severe illness among high-risk groups such as elderly and immunocompromised patients. Mandatory influenza vaccination of healthcare workers (HCWs) has been viewed as means to improve patient and HCW protection.

**Methods:** We analyzed data collected by a web-based survey sent annually to all Finnish acute care hospitals and described the influenza vaccination coverage among HCWs during seasons 2017/18 (without mandate) and 2018/19 (mandate enforced).

**Results:** In season 2017/2018, 38/39 hospitals provided data and 35/36 hospitals in 2018/2019. The mean coverage in season 2017/18 was 88% (median, 84%; range 48-100%) and in 2018/19 91% (median, 91%; range 57-100%). The mean increase from season 2017/2018 to 2018/2019 was 6.5% (median, 3%; range -11.0-33.0%).

**Conclusions:** The coverage of influenza vaccinated HCWs in Finnish hospitals was high. However, there were major differences between hospitals which raise the question about data quality as well as implementation of the mandate, and need further evaluation.

## Introduction

Influenza can cause severe illness among high risk groups such as the elderly and immunocompromised patients, leading to several million severe cases of illness and between 250,000 and 500,000 deaths annually (1). The primary prevention for influenza is vaccination.

Higher influenza vaccination coverage among health care workers (HCW) is associated with lower mortality of patients and also reduced staff sick days, leading to better care provision (2-5).

However, among HCWs worldwide, vaccination coverage is unsatisfactory (6, 7). The barriers to vaccination among HCWs include lack of knowledge about both disease and vaccine, doubts about the safety and the effectiveness of the vaccine, dislike of injections, fear of adverse effects, inconvenience, and limited support from managers (8, 9). Among nurses, knowledge and risk perceptions are the strongest predictors of vaccination uptake. (10) In order to improve the design and delivery of interventions, information about influenza vaccination coverage among HCWs is essential.

To improve immunisation uptake and protect patients, Finland recently introduced the National Communicable Diseases Act (12), which mandates influenza vaccination for HCWs, some social workers, and students in health care professions. The Act entered into force on 1 March 2017, with the relevant section (Section 48) coming into force one year later.

Almost all European countries recommend influenza vaccination for HCWs, however influenza vaccine uptake can be as low as 40% (13). We are not aware of any European countries that mandate influenza vaccination for HCWs except Finland. We investigated the impact of the new Act by measuring immunisation uptake in the 2018/2019 influenza season, the first season after the change in the legal basis, and by comparing it to uptake in the previous season.

## Methods

The healthcare system in Finland is organised into 20 geographically and administratively defined healthcare districts (HDs), with populations ranging from 63 000 to 1.8 million. Fifteen HDs have only secondary and other care hospitals, and five also provide tertiary care services. HDs have a consulting role in the infection control activities in their area.

Starting at the beginning of 2018 a web-based survey requesting data on structure and process indicators related to infection control has been sent annually to all Finnish acute care hospitals. Psychiatric and long-term care facilities were excluded from the survey, as well as hospitals on the Åland Islands. One of the survey questions concerns influenza vaccination coverage among HCWs. The survey was sent to staff who are practically in charge of the infection control activities. During 2018-2019, all five tertiary care hospitals, 15 secondary care hospitals and 19 other hospitals participated in the survey.

The unit of analysis in the study was hospitals, as only aggregate data (vaccination coverage per hospital) were available. Aggregation was done at hospital level by using the following formula and specifications (14):

- The numerator is the total number of vaccinated HCWs in *special health care units on December 31. HCWs include nurses and physicians, but not administrative and technical staff*.
- The denominator is the total number of HCWs in special health care units at a given time, for example on Monday of week 44. Offsets are limited to those who were absent for more than 30 days on that day, regardless of the reason. HCWs includes nurses and physicians, but not administrative and technical staff.
- Depending on the framework, psychiatry may be considered, for example, if it is located in the same building as the somatic specialist wards. Laboratory staff will be taken into account if readily available. Most importantly, the numerator and denominator data in the formula should cover personnel in the same units / specialties.

The main indicator used in the study was: influenza vaccination coverage among HCWs in percent per hospital and HD. The data analysis consisted of the description of the distribution of main indicators. All analysis was done in MS Excel.

## Results

Out of the 39 hospitals that provided data on influenza vaccination coverage among HCW, 34 provided data for both seasons. In the first season, 38 hospitals (survey sent to 39 hospitals) reported the coverage and 35 (survey sent to 36 hospitals) in the second season. The coverage and changes in coverage differed across individual hospitals and HDs (table 1). In season 2017/18, the mean coverage in hospitals was 88% (median, 84%; range by hospitals, 48-100%) and 91% (median, 91%; range by hospitals, 57-100%) in season 2018/19. Immunisation uptake increased from season 2017/18 to season 2018/2019 by 6.5% (median, 3.0%; range by hospitals, -11.0-33.0%). In season 2017/18, 12 hospitals had a coverage of 90% or above and 2 hospitals reached 100% while a year after 22 hospitals reported a coverage of 90% or more and 8 hospitals reached 100%.

**Table 1.**
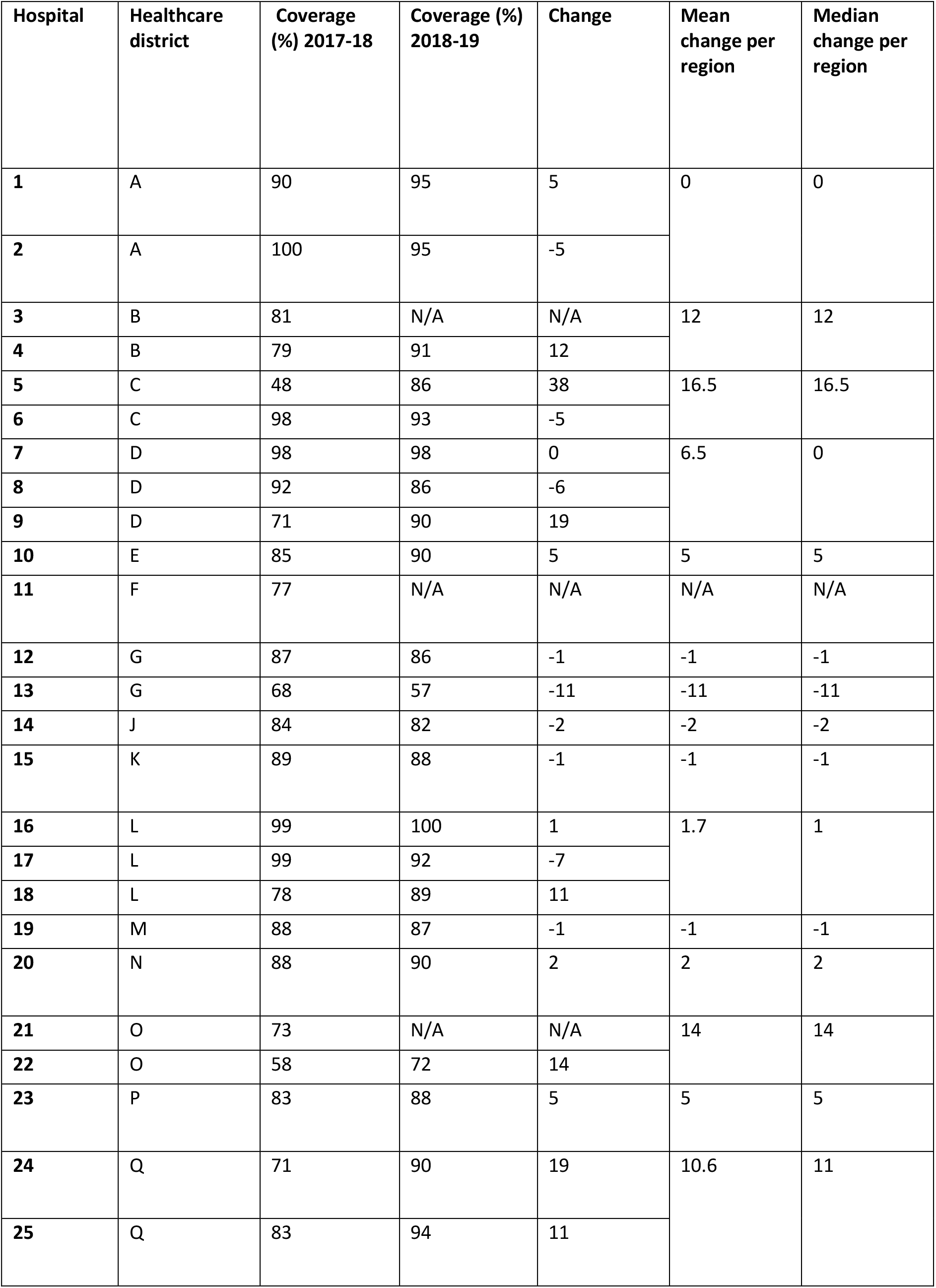
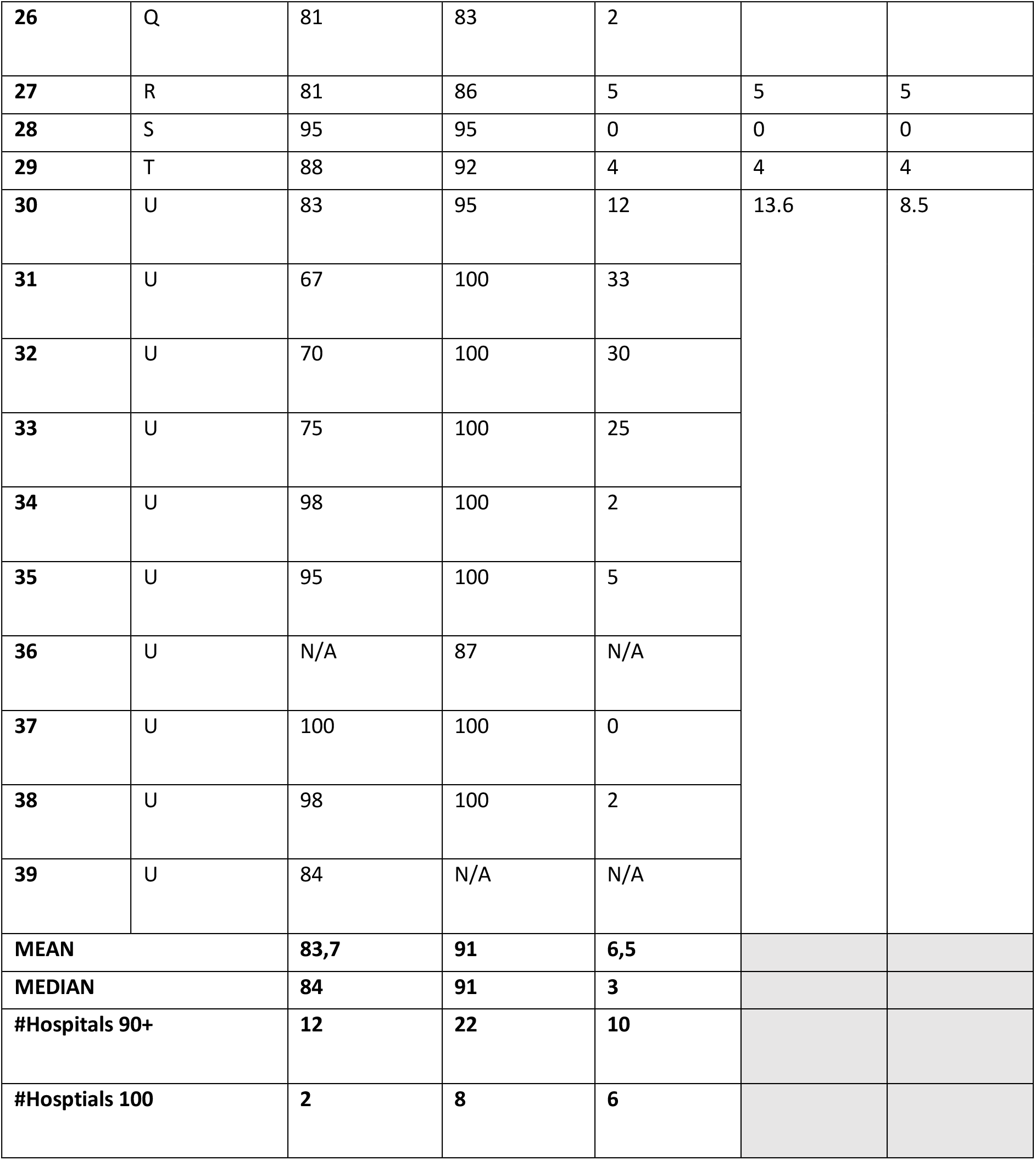
Influenza vaccination coverage among healthcare workers during seasons 2017/2018 and 2018/2019 with change by hospital and healthcare district

Figure 1 shows the coverage per HD during both seasons. For 11 HDs the coverage increased, for 5 it decreased, for 2 it remained the same and for 1 there was too much data missing to estimate the change in HCW influenza vaccination coverage (figure 2).

**Figure 1.**
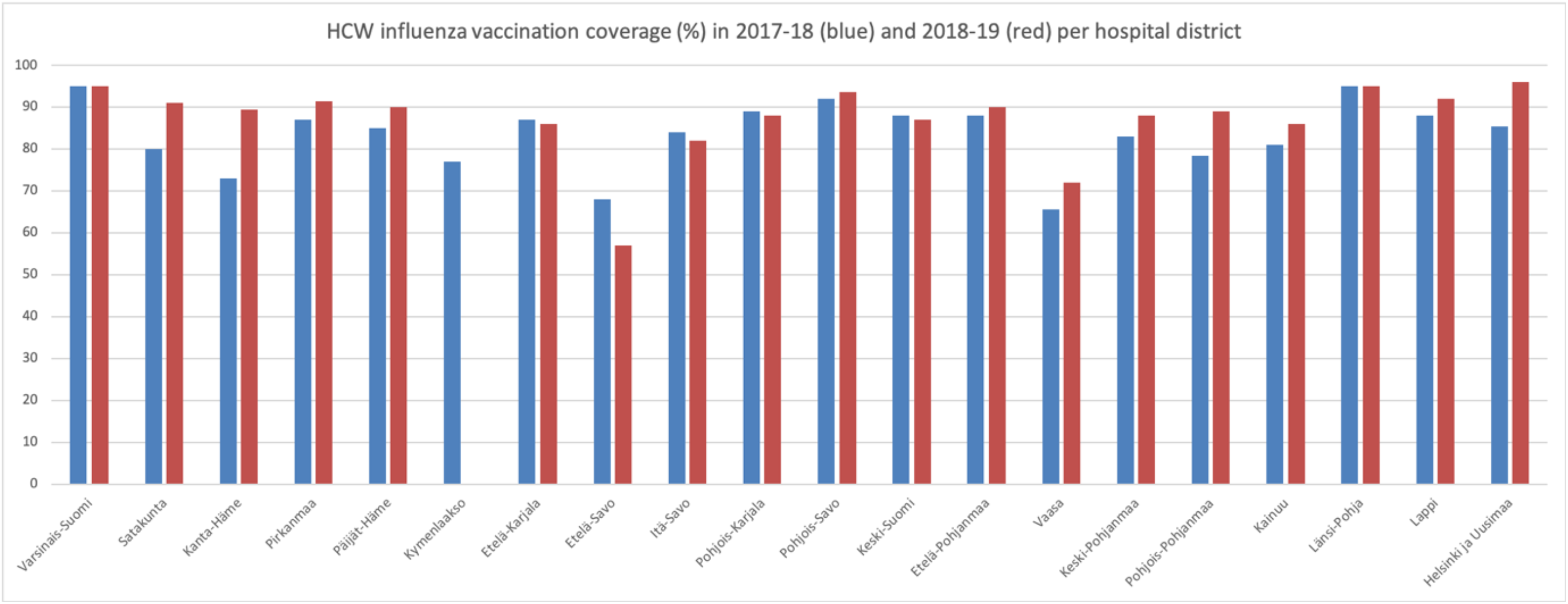
Influenza vaccination coverage (%) among healthcare workers during seasons 2017/18 and 2018/19 by healthcare district

**Figure 2:**
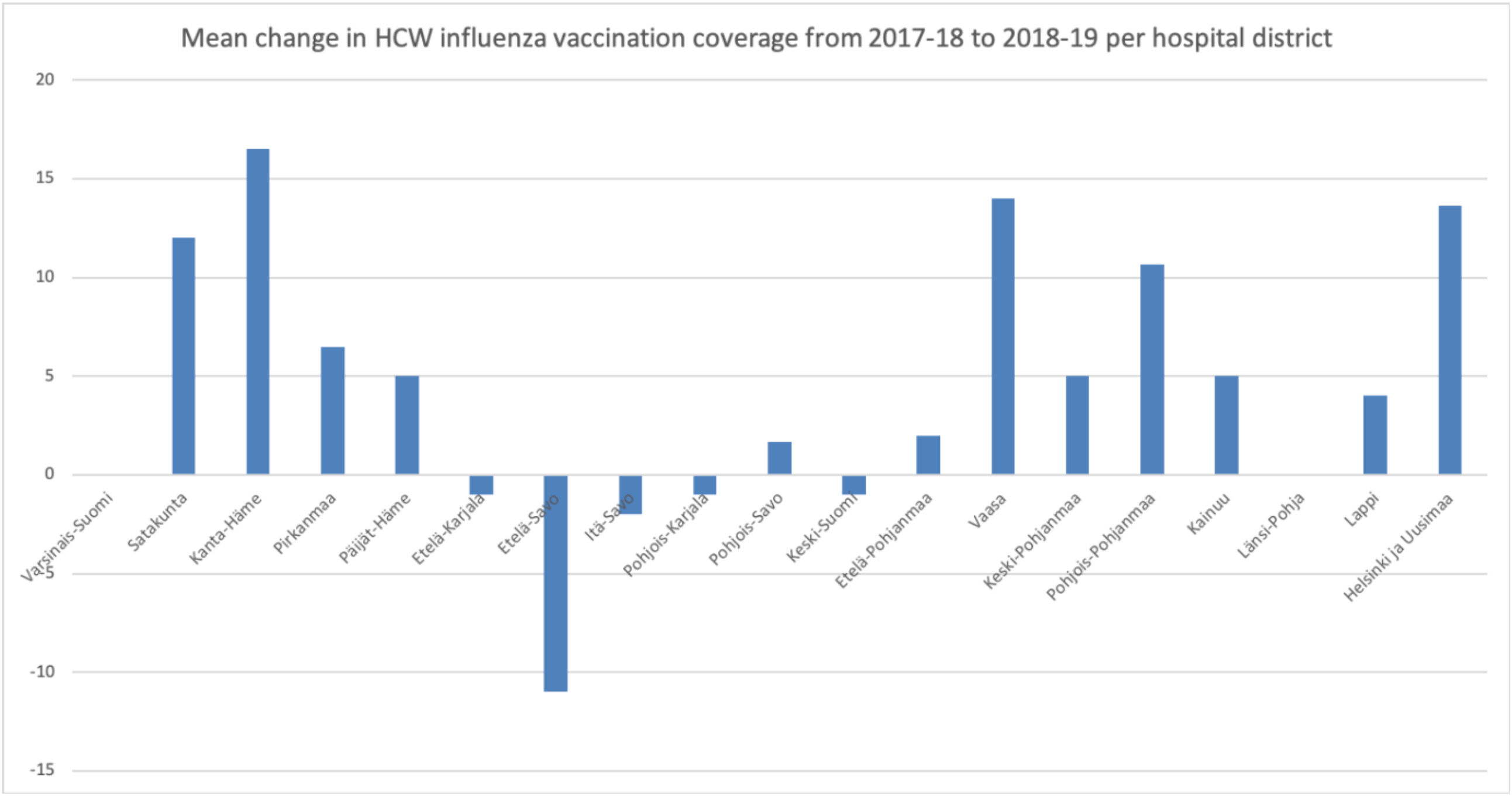
Mean change in influenza vaccination coverage among healthcare workers from season 2017/18 to season 2018/19 by healthcare district

## Discussion

Generally speaking, the percentage of vaccinated HCWs in Finland was high with a hospital mean of 88% in 2017/2018 and of 91% in 2018/2019. In comparison, percentages in the US ranged from 77% to 81% over the seasons 2014/15 to 2018/19 (15) and in England from 63% to 70% in the seasons 2016/17 to 2018/19 (16, 17). As uptake was already relatively high, the introduction of the mandate in the Communicable Disease Act probably had less of an effect than it might have in other countries. However, it appeared to have an effect in bringing those hospitals and HDs that were previously underperforming up to a higher coverage as well as by raising the number of hospitals with 100% vaccinated HCWs from two to eight. As no further data are available, the interpretation of the results and especially the comparison across hospitals and HD is uncertain. However, the sometimes-applied term ‘semi-mandatory’ seems to be suitable for the provisions made in the 2016 Finnish Communicable Diseases Act.

The method for data collection used in this case warrants some further evaluation as it differs from approaches taken in other countries. The current Finnish method is highly decentralised, with calculations of the percentage being performed in each hospital and only percentage data being reported to the National Public Health Institute (Finnish Institute for Health and Welfare, THL). To our knowledge, many hospitals use one specific commercial software in the data collection. By comparison, the system in England relies on Trusts, which are similar to HDs in Finland for data collection, however, percentages are calculated centrally with each Trust reporting cumulative numbers of influenza vaccine doses given to HCWs (18). Additionally, data in England is collected in five monthly surveys rather than once yearly (18). This also allows for analysis of uptake throughout the influenza season, which the Finnish system does not allow. On the other end of the methodological spectrum, the US system uses self-reported vaccination uptake through an online panel (15, 19). The reliability of this approach, however, being unclear. The US system has been used in previous years and also includes weighting to the distribution of the US population of healthcare personnel by occupation, age, sex, race/ethnicity, work setting, and U.S. Census region (20). While this does not give a probability sample, and the Centers for Disease Control and Prevention (CDC), therefore, do not perform any statistical analyses with the data, it provides the added benefit of not relying on HDs or similar entities, all of which have their own vested interests, in the collection of the data. However, the data collection done by the CDC happens in a very different setting and health care system and conveniences and adjustments in methodology that are needed in such a system are not necessary on the Finnish case, where public health care is the standard. The finer details, such as the weighting, could, however, be considered further in refining the system in Finland.

This study is subject to a number of potential biases and limitations, the first of which being the availability of percentages of vaccinated staff only. As hospitals calculate the vaccination coverage among their HCW themselves there is a possibility for error and differences in calculation across hospitals/HDs. While there is an official suggested formula for hospitals to use, there is no way for ensuring this was done and done correctly. Additionally, we cannot rule out different data collection in different hospitals/HDs. There is a risk that hospitals and HDs use different data and measures for their calculation of vaccination coverage (e.g. for denominator staff numbers at different times of the year). An additional major limitation is that no data on influenza vaccination coverage among HCWs was systematically collected before 2017 in Finland. Finally, it can be assumed that at least for some hospitals reaching 100% this is due to a too small denominator, which is difficult to estimate correctly due to changes in staff and other logistical issues. Hence, it is difficult to conclude whether the observed changes are part of a larger trend and what the exact contribution of the mandate was.

## Conclusion

Influenza vaccination coverage among HCWs in Finland is high and possibly increasing. While some HDs are performing better than others in this regard the overall coverage is encouraging. While the analysis of just two seasons does not provide sufficient evidence for an overall trend the changes observed across the two seasons seem positive as well.

The semi-mandatory system that Finland is employing can be seen as a compromise to navigate the moral dilemma of pitting the interest and rights of the individual HCW against to interests and rights of patients. This is particularly the case as the law leaves latitude for both organisations and individuals to not only make administrative decisions, such as hiring unvaccinated HCWs because no vaccinated HCWs are locally available at the time, but also moral ones. This obviously leads to potential questions for further inquiry regarding the intention of the vagueness of the law, the different interpretations of the law by HCWs, hospital administrators and municipalities as well as detailed legal analysis.

Regarding the data available at the moment, there is room for improvement. Despite the unclear data quality at the moment, a first important recommendation in both the Finnish context but also more generally is to keep collecting this type of data. The need for analysis like this after changes in the context shows that routine data collection regarding HCWs influenza vaccination coverage are essential not only for routine decision making but also for assessing the impact of changes such as the ones in Finland. However, the quality of the data should also be improved. HDs reporting of crude data would be more advisable so that percentages can be calculated centrally with an ensured standardised methodology. Additionally, it should be further investigated if weighting to the distribution of the population of healthcare personnel by occupation, age, sex, race/ethnicity, work setting, and region (such as done in the US context) might yield additional useful information. This might most suitably be done in the form of an initial pilot study in two or three hospital.

Finally, beyond the additional routine data collection and research suggested above, further analysis of the legal context could provide deeper insights and it would also be beneficial after changes to the legal context like this to assess the reception of the new law in the main target group. For this purpose, it would be beneficial to study how HCWs’ and hospitals’ attitudes to influenza vaccination have been influenced by the law and what their knowledge, attitudes and practices regarding the new legal basis for ‘semi-mandatory’ influenza vaccination due to their profession are.

## Data Availability

The disaggregate data is confidential.

